# How were Hospitals Affected by the Ministry’s Release of Hospital Names to be Potentially Reorganized?

**DOI:** 10.1101/2024.02.27.24302544

**Authors:** Hiromichi Takahashi, Jung-ho Shin, Susumu Kunisawa, Kiyohide Fushimi, Yuichi Imanaka

## Abstract

**Background:** The Japanese Ministry of Health, Labour and Welfare (MHLW) released a list of public and municipal hospitals (hereinafter “the list”) that are subject to reevaluation for hospital function. First, this study describes the functional differentiation status of Japanese hospital beds. Second, it evaluates the impact of the list release on the number of admissions in the listed hospitals.

**Methods:** Firstly, the number of hospitals and beds by the function of listed and non-listed hospitals in 2019 and 2021 were described using the bed function report. The Controlled Interrupted Time Series (CITS) analyses were subsequently conducted using Diagnosis Procedure Combination (DPC) data. Hospitals were divided near the cutoff point of the list. The outcomes were the number of admissions for gastrointestinal cancer surgery, those admitted via ambulance, or with a femoral fracture per 1,000 admissions. The exposure point was the week when the list was released.

**Results:** A decrease in the total number of beds was observed in 18.9% of the listed hospitals and 10.2% of others. Changes in bed functions were observed in 19.9% of the listed hospitals and 12.5% of others. CITS analyses showed that the rate ratio of admissions for gastrointestinal cancer surgery, those admitted via ambulance, and those with a femoral fracture in the listed hospital group after the list’s release were 1.001 (95% CI: 0.998–1.004, p = 0.619), 1.001 (95% CI: 0.998–1.004, p = 0.548), and 0.998 (95% CI: 0.998–1.002, p = 0.313), respectively.

**Conclusion:** More prominent trends of functional differentiation of hospital beds were observed in the listed hospitals. The release of the list did not impact the number of hospital admissions for gastrointestinal cancer surgery, those admitted via ambulance, or those with a femoral fracture per 1,000 admissions in the listed hospitals.

## 1. Introduction

Japan has the most aged population in the world, and the accompanying imbalance between healthcare supply and demand is a challenge. The number of elderly people aged 75 and over will likely exceed 20 million by 2025,^1–3^ and this aging population is changing healthcare demands.^4^ The importance of post-acute and convalescent care is increasing, as the elderly require more recovery time than younger patients. In contrast, after World War II, the Japanese healthcare delivery system focused mainly on acute care.^4–6^ A balanced healthcare delivery system with adequate supply and demand is required in Japan.

To address this imbalance and achieve an optimized healthcare delivery system, the Ministry of Health, Labour and Welfare (MHLW) introduced a Community Health Care Vision (CHCV) in 2016.^7^ The CHCV includes the necessary number of beds by healthcare function in 2025 and the actual number of beds by healthcare function chosen by each hospital for each of the 341 vision areas nationwide. Under the system of reporting on the bed functions,^8^ each hospital must report its present and future (i.e., 2025) medical functions by choosing from the following: advanced acute care, acute care, convalescent, and chronic care.^9^ A CHCV coordination meeting was established for each vision area,^9, 10^ composed of representatives of community medical associations, dental associations, pharmaceutical associations, nursing associations, municipalities, medical insurers, healthcare providers, and others. Currently, there is an oversupply and undersupply of acute care and convalescent beds, respectively, compared to the demand for such beds. Under the CHCV, each hospital is expected to promote functional differentiation by voluntarily selecting the functions of its own beds based on stakeholder discussions within the vision area.

Despite introducing the CHCV, the functional differentiation of hospital beds was not promoted easily. It was pointed out that the CHCV and its coordination meeting had contributed little to the differentiation of hospital bed functions.^2, 11, 12^ One reason for this is that the Japanese healthcare system depends greatly on the private sector — 68.2% of all hospitals were owned privately in 2016 ^13^ — and healthcare providers have considerable freedom to choose hospital functions, making it difficult for the MHLW to promote reform.^14^ Thus, the MHLW attempted to start with reforming public and municipal hospitals, which are relatively controllable. The CHCV Working Group led by the MHLW analyzed data on clinical performance related to the acute phase functions of public, municipal and some private hospitals serving public utilities in response to the Basic Policy on Economic and Fiscal Management and Reform, 2019 and 2018.^15, 16^ As a result, MHLW designated 424 out of 1,455 public and municipality hospitals as “hospitals that need further consideration regarding the necessity of reorganization and integration” (hospitals subject to reevaluation, hereinafter, “listed hospitals”). It released a list with the names of these hospitals and their medical performance data (hereinafter, “the list”) on September 26, 2019.^11^ According to MHLW, reorganization and integration include downsizing, functional differentiation, and consolidation.^17^ Moreover, the number of hospital beds by function was the main monitoring indicator before and after the publication of the list^18^ and subsidies for reorganization also encourage functional differentiation and reduction of hospital beds.^19^ As such, the publication of this list was mainly focused on functional differentiation of hospital beds. MHLW established two selection criteria regarding the designation of the listed hospitals: that they “have only a small number of medical practices” and are “similar and close to others.” MHLW set Categories A and B, including nine and six evaluation items, respectively.^20^ If a hospital has fewer cases of all nine items in Category A than the number of cases corresponding to the 33.3 percentile among all the public and municipality hospitals, the hospital meets the former criteria (“has only a small number of medical practices”). On the other hand, if there are two or more hospitals that have a high number of cases for all six items among Category B in the same vision area,^9, 10^ and each hospital is located less than 20 minutes away by car, hospitals in this vision area meet the latter criteria (“similar and close to others”). Hospitals were designated as the listed hospitals if they met either of two criteria. The MHLW declared that the release of the list aimed to stimulate discussions at the CHCV coordination meetings based on the current and future regional visions. Despite the MHLW’s intent, the media described the list as a “hospital restructuring list” forced by the government.^21^ Furthermore, the selection criteria, evaluation items, and analysis methods were pointed out as biased and arbitrary.^22, 23^ The National Governors’ Association and the Japan Medical Association expressed concern that the list would cause chaos in local communities.^24–27^

The policy effects of this list publication have not been adequately objectively discussed. The MHLW reported that progress in functional differentiation has been made as of 2021 for the hospitals listed in 2019.^18^ From 2019 to 2021, the total number of beds in the listed hospitals decreased by approximately 4,000, including a decrease of approximately 4,100 acute care beds and an increase of approximately 1,700 convalescent beds.^18^ However, whether this functional differentiation differs from that in non-listed hospitals is unclear, and the impact of the publication of the list on factors other than the differentiation of hospital bed functions requires study. Even if the number of beds by function did not change, progress can be thought made if it promotes discussion and role-sharing in the community, and if reputational damage becomes apparent, it should be corrected. Therefore, two objectives were set in this study. The first objective was to clarify the status of functional differentiation of hospital beds in listed hospitals compared to non-listed hospitals in Japan. The second objective was to evaluate the impact of the release of the list on the number of admission cases at the listed hospitals. The admission cases analyzed were admissions for gastrointestinal cancer surgery, admissions via ambulance, and admissions with a femoral fracture, which are included in the evaluation items in Categories A and B when the listed hospitals were selected.^11^

## 2. Materials and Methods

### 2.1. Description of functional differentiation of hospital bed status of listed hospitals

The data source used was the report on the bed functions released annually by the MHLW.^8^ The bed function reporting system requires hospitals nationwide to report based on hospital wards starting in FY 2014. The report on the bed functions system covers only licensed general and long-term care beds defined in the Medical Service Act,^8^ hereinafter, “Licensed beds”; this includes general and long-term care beds, but not psychiatric, infectious disease, or tuberculosis beds. The hospitals having general or long-term care beds are required to report on the number of beds by functions, number of patients, number of workforce, number of medical devices, number of surgeries, and other specific medical details, with approximately 400 items of data as of July 1 of each year.

Hospitals included in the report in 2017 were classified into two classes: listed and non-listed. Listed hospitals were 417 hospitals out of the 424 listed hospitals initially selected for reevaluation^11^, excluding the 7 later exempted from the list.^28^ For both listed and non-listed hospitals, those that did not report on the number of beds by function in either 2019 or 2021 were excluded. Listed and non-listed hospitals that reported the number of beds by function in 2019 and 2021 were classified into three classes: those that increased, decreased, or saw no change in the total number of licensed beds from 2019 to 2021. Hospitals with no change in the total number of licensed beds, were furthermore grouped into those with and those without a change in the number of licensed beds by function, i.e., advanced acute care phase, acute care phase, convalescent phase, chronic care phase, and inactive. The number of hospitals in each classification is described. In addition, the total number of licensed beds in 2019 and 2021, the proportion of change in the number of beds, and the change in the number of beds per hospital in the same period were described by each function.

### 2.2. Impact of the release of the list on admissions to the listed hospitals

#### Data source and study population

The Diagnosis Procedure Combination (DPC) database of a research group funded by the MHLW was used for this study. The database consists of DPC data from mainly acute care hospitals that are voluntarily participating. Approximately 1,200 hospitals are participating across Japan, comprising both public and private hospitals.^29^ The DPC/per-diem payment system (PDPS) is a Japanese prospective payment system applied to acute care hospitals. Although not all DPC/PDPS hospitals are included in the DPC research database, individual data are available for research purposes^30^. As described in detail elsewhere,^31^ the DPC data includes insurance claims, the patient’s clinical characteristics, and records of clinical services. The clinical characteristics include age, gender, admission and discharge statuses and date, cause of admission, primary diagnosis, diagnosis for which most and second most of the medical resources, and other data. All diagnoses were classified according to the International Classification of Diseases, 10th Revision (ICD-10) codes.

Among the research database, the study included public and municipal hospitals in vision areas with a population of less than 1 million and those provided with DPC data Form 1 for 36 months from April 2018 to March 2021. Hospitals in vision areas with a population of 1 million or more were excluded because public and municipal hospitals in such areas were not requested to reevaluate even if they met the “similar and close to others” criterion.^17^ The selected hospitals were divided into the listed hospital group and the controlled hospital group. The listed hospital group consisted of hospitals that met the criteria of the 417 listed hospitals in Section 2.1. The control hospital group consisted of hospitals that were not included in the listed hospitals and met eight of the nine evaluation items in Category A or five of the six evaluation items in Category B. The individual hospitals around this cutoff in both listed hospital group and controlled hospital group are almost homogeneous, considered to “have only a small number of medical practices” or “similar and close to others.” The difference between the two groups is considered to be whether or not they were selected as listed hospitals. By comparing outcomes between the two groups, it is possible to consider the observed difference to be the impact of being selected as listed hospitals.

The study period lasted 143 weeks, from week 14, 2018 (week beginning Monday, April 2, according to ISO 8601) to week 52, 2020 (beginning December 21). While the inclusion hospitals were those that provided 36 months of DPC data, the reason for setting the study period as 143 weeks (about 33 months) was that DPC data is generated monthly after discharge from the hospital. If the study period is set up to the end of March 2021, DPC data for cases admitted by the end of March 2021 and discharged after April 2021 will not be generated, resulting in an undercount of the actual number of cases.

Three types of cases newly admitted to the listed and control hospital groups during the study period were extracted: those who underwent gastrointestinal cancer surgery, those arriving via ambulance, and those with a femoral fracture. These three case types were identified as the target population. The process for selecting the three types of cases had three steps. Firstly, the three types of cases were selected among evaluation items included in Categories A and B. The number of cases (for only June every year) in each evaluation item included in Categories A and B is disclosed and is considered to be likely affected by the release of the list. Secondly, among all the nine and six evaluation items in Categories A and B, respectively, the evaluation items that rely on facilities and specialized personnel, such as cardiac, pediatric, and obstetric care, were excluded because the short-term effects of the release of the list are not expected. Finally, to examine the impact of the release of the list on major types of cases, the cases that were not treated by more than 70% (calculated using the list) of the listed hospitals were excluded in this study,^11^. Admission cases for gastrointestinal cancer surgery were identified based on the following conditions: “Name of disease with the most resource-intensive” in DPC data was an ICD-10 code for gastrointestinal cancer, and the corresponding surgery was performed (Supplemental Table 1). Admission cases via ambulance were identified as having a check of “Ambulance transport” in DPC data. Admission cases with a femoral fracture were identified as having ICD-10 code S72.x for either “Name of trigger diagnosis causing admission” or “Name of disease with the top input of medical resources” in DPC data. Admission cases missing “ambulance transport”, “scheduled/emergency admission classification”, or “discharge outcome” in Form 1 were excluded from the analysis.

#### Statistical analysis

For each of the listed hospital groups and the control hospital group, we described the establishment entity (the national government, public organizations, social insurance bodies, medical corporations, or others),^32^ the number of beds by function (total, advanced acute care, acute care, convalescent, chronic care, and inactive), region (Hokkaido, Tohoku, Kanto, Chubu, Kinki, Chushikoku, or Kyusyu), vision area category by population size (500,000–1 million, 200,000–500,000, 100,000–200,000, or less than 100,000), whether the hospital is a special functioning hospital or a regional medical care support hospital,^10, 33^ and the status of emergency hospital designation by MHLW (primary, secondary, or tertiary emergency), reported in the report on the bed functions 2019. For each of the three types of admission cases, age, gender, and length of stay in the hospital were described before and after the release of the list for each listed hospital group and each control hospital group.

Controlled Interrupted Time Series^34^ (CITS) analysis was performed to estimate the impact of the release of the list on the number of three types of admission cases in all the listed hospital group compared to the controlled hospital group. The dependent variables were the number of admissions for each of the three types of admissions divided by the number of 1,000 new admissions to investigate the possibility that patients, ambulance teams, and other medical institutions may not choose the listed hospitals after the list was released, or that the listed hospital may have restructured its functions in response to the release of the list that showed the low number of cases. Poisson or quasi-Poisson distributions were assumed considering the data’s overdispersion. Seasonality was considered by including harmonic terms.^35^ The 39th week of 2019 (beginning September 23) was considered the exposure week, including September 26, 2019, the day of the list’s release at the 24th working group meeting on the CHCV. Although the list was released on September 26, 2019, the MHLW officially requested each prefecture to reevaluate the listed hospitals’ functions on January 17, 2020. To account for the possibility of the impact on listed hospitals by triggering the official request, a sensitivity analysis where exposure week was the 3^rd^ week of 2020 was performed. The model used was explained in detail in Appendix. Our primary interest was the difference in the slope change between the listed hospital group and the control hospital group.

A two-sided p-value of 5% was considered statistically significant. CITS analysis was performed using R (R Foundation for Statistical Computing, Vienna, Austria) version 4.2.1 and tsModel package version 0.6.1. Data used in this study were anonymized and approved by the Ethics Committee, Graduate School of Medicine, Kyoto University (Approval No.: R0135).

## 3. Results

### 3.1. Description of functional differentiation of hospital bed status in listed hospitals

Some 7,244 hospitals included in the report on the bed functions in FY 2017 were classified into 417 listed hospitals and 6,827 non-listed hospitals. The number of listed and non-listed hospitals that reported the number of licensed beds in both 2019 and 2021 were 398 and 6,415, respectively (Supplemental Figure 1). Table 1 shows the functional differentiation classification of listed and non-listed hospitals. Differences were observed between the listed and non-listed hospitals in the decrease in the total number of licensed beds (χ^2^(1) = 31.92, p < 0.01) and change in the number of licensed beds by function (χ^2^(1) =19.08, p < 0.01). For 2019 and 2021, Table 2 shows the number of licensed beds, the proportion of change in the number of licensed beds, and the mean change in the number of beds per hospital by function for each of the listed and non-listed hospitals that reported the number of licensed beds in both 2019 and 2021.

**Table 1.** Description of functional differentiation status (n, (%))

|  | Listed hospitals (n = 398) |  | Non-listed hospitals<br>(n = 6,415) |  |
| --- | --- | --- | --- | --- |
| Increase in the total number of beds | 14 | ( 3.4) | 411 | ( 6.0) |
| Decrease in the total number of beds | 79 | ( 18.9) | 693 | ( 10.2) |
| No change in the total number of beds |  |  |  |  |
| with change in the number of beds by function | 83 | ( 19.9) | 854 | ( 12.5) |
| without change in the number of beds by function | 222 | ( 53.2) | 4,457 | ( 65.3) |

**Table 2.** Number in 2019 and 2021, change proportion, and change in number per hospital of beds by function.

|  |  | Total | Advanced<br>acute care | Acute care | Convalescent | Chronic care | Inactive |
| --- | --- | --- | --- | --- | --- | --- | --- |
| Listed hospitals<br>(n = 398) | Number of beds in 2019 | 62,693 | 1,252 | 32,681 | 13,312 | 12,940 | 2,508 |
|  | Number of beds in 2021 | 61,099 | 1,100 | 30,027 | 15,655 | 12,489 | 1,828 |
|  | Change proportion | -2.5% | -12.1% | -8.1% | 17.6% | -3.5% | -27.1% |
|  | Mean change in the number of<br>beds per hospital | -4.01 | -0.38 | -6.67 | 5.89 | -1.13 | -1.71 |
| Non-listed hospitals<br>(n = 6,415) | Number of beds in 2019 | 1,091,582 | 155,968 | 466,307 | 154,742 | 294,998 | 19,567 |
|  | Number of beds in 2021 | 1,078,083 | 152,772 | 469,166 | 159,687 | 280,739 | 15,719 |
|  | Change proportion | -1.2% | -2.0% | 0.6% | 3.2% | -4.8% | -19.7% |
|  | Mean change in the number of<br>beds per hospital | -2.10 | -0.50 | 0.45 | 0.77 | -2.22 | -0.60 |

### 3.2. Impact of the release of the list on the admission to the listed hospitals

Among the hospitals included in the DPC research database, 90 hospitals were extracted that met the criteria described previously: public and municipal hospitals in vision areas with a population of less than 1 million and those provided with DPC data Form 1 for 36 months from April 2018 to March 2021. The entire number of cases admitted to these hospitals during the 143-week study period (April 2, 2018–December 27, 2020) was 1,148,442. The 90 extracted hospitals comprised 40 hospitals in the listed hospital group and 56 hospitals in the control hospital group, with total admissions of 369,692 and 778,750 cases, respectively. Table 3 shows the characteristics of hospitals in each group. Table 4 shows the patients’ background of each case before and after the release of the list.

**Table 3.** Characteristics of hospitals in each group.

|  | Listed hospital group |  | Control hospital |  |
| --- | --- | --- | --- | --- |
|  | (n = 40) |  | group (n = 56) |  |
| **Region, n (%)** |  |  |  |  |
| Hokkaido | 2 | (5.0) | 1 | (1.8) |
| Tohoku | 3 | (7.5) | 4 | (7.1) |
| Kanto | 7 | (17.5) | 10 | (17.9) |
| Chubu | 13 | (32.5) | 16 | (28.6) |
| Kinki | 4 | (10.0) | 6 | (10.7) |
| Chushikoku | 7 | (17.5) | 11 | (19.6) |
| Kyusyu | 4 | (10.0) | 8 | (14.3) |
| **Vision Area category by population, n (%)** |  |  |  |  |
| 500,000–1 million | 22 | (55.0) | 27 | (48.2) |
| 200,000–500,000 | 17 | (42.5) | 20 | (35.7) |
| 100,000–200,000 | 1 | (2.5) | 4 | (7.1) |
| Less than 100,000 | 0 | (0.0) | 5 | (8.9) |
| **Emergency hospitals, n (%)** |  |  |  |  |
| Tertiary emergency facility | 0 | (0.0) | 2 | (3.6) |
| Secondary emergency facility | 35 | (87.5) | 50 | (89.3) |
| Primary emergency facility | 37 | (92.5) | 52 | (92.9) |
| **Approval status, n (%)** |  |  |  |  |
| Special functioning hospital | 0 | (0.0) | 0 | (0.0) |
| Regional medical care support hospital | 12 | (30.0) | 27 | (48.2) |
| **Establishment entity, n (%)** |  |  |  |  |
| National government | 8 | (20.0) | 23 | (41.1) |
| Public organizations | 25 | (62.5) | 24 | (42.9) |
| Social insurance bodies | 2 | (5.0) | 2 | (3.6) |
| Medical corporations | 1 | (2.5) | 3 | (5.4) |
| Others | 4 | (10.0) | 4 | (7.1) |
| **Number of beds by function, median (IQR)** |  |  |  |  |
| Total | 226.5 | (180, 308) | 291 | (236, 369) |
| Advanced acute care | 0 | (0, 0) | 3.5 | (0, 12) |
| Acute care | 149 | (120, 215) | 175 | (118, 256) |
| Convalescent | 47 | (0, 60) | 48 | (0, 75) |
| Chronic care | 0 | (0, 52) | 0 | (0, 38) |
| Inactive | 0 | (0, 0) | 0 | (0, 3) |

**Table 4.** Patient background of each case.

|  | Listed hospital group (n = 40) |  |  |  | Control hospital group (n = 56) |  |  |  |
| --- | --- | --- | --- | --- | --- | --- | --- | --- |
|  | Before the release |  | After the release |  | Before the release |  | After the release |  |
|  | of the list |  | of the list |  | of the list |  | of the list |  |
| Admission cases for gastrointestinal cancer surgery |  |  |  |  |  |  |  |  |
| Number of cases, n | 6,063 |  | 5,077 |  | 15,440 |  | 12,724 |  |
| Gender - male, n (%) | 3,736 | (61.6) | 3,158 | (62.2) | 9,751 | (63.2) | 8,073 | (63.5) |
| Age, median (IQR) | 74 | (67, 81) | 74 | (68, 81) | 72 | (65, 79) | 72 | (65, 79) |
| Length of stay, median (IQR) | 17 | (10, 31) | 16 | (9, 29) | 14 | (8, 24) | 13 | (8, 22) |
| Admission cases via ambulance transport |  |  |  |  |  |  |  |  |
| Number of cases, n | 33,285 |  | 26,919 |  | 67,584 |  | 53,276 |  |
| Gender - male, n (%) | 16,336 | (49.1) | 13,299 | (49.4) | 34,834 | (51.5) | 27,497 | (51.6) |
| Age, median (IQR) | 80 | (68, 87) | 80 | (70, 88) | 78 | (65, 86) | 79 | (67, 87) |
| Length of stay, median (IQR) | 13 | (5, 31) | 14 | (5, 32) | 13 | (5, 28) | 14 | (6, 28) |
| Admission cases with a femoral fracture |  |  |  |  |  |  |  |  |
| Number of cases, n | 4,097 |  | 3,552 |  | 6,196 |  | 5,315 |  |
| Gender - male, n (%) | 922 | (22.5) | 817 | (23.0) | 1,513 | (24.4) | 1,256 | (23.6) |
| Age, median (IQR) | 85 | (78, 90) | 85 | (78, 90) | 85 | (77, 90) | 85 | (78, 90) |
| Length of stay, median (IQR) | 42 | (24, 64) | 39 | (23, 59) | 32 | (20, 56) | 30 | (20, 53) |

\* Before and after the release of the list indicate the period from April 2, 2018, to September 25, 2019, and from September 26, 2019, to December 27, 2020, respectively.

Figure 1 shows the weekly outcomes by case in each listed and controlled hospital group with the result of the CITS analysis. The results of the CITS analysis showed that the impact of the release of the list on the outcomes in the listed hospital group was not statistically significant compared to one in the control hospital group, both in the primary and sensitivity analyses.

**Figure 1.**
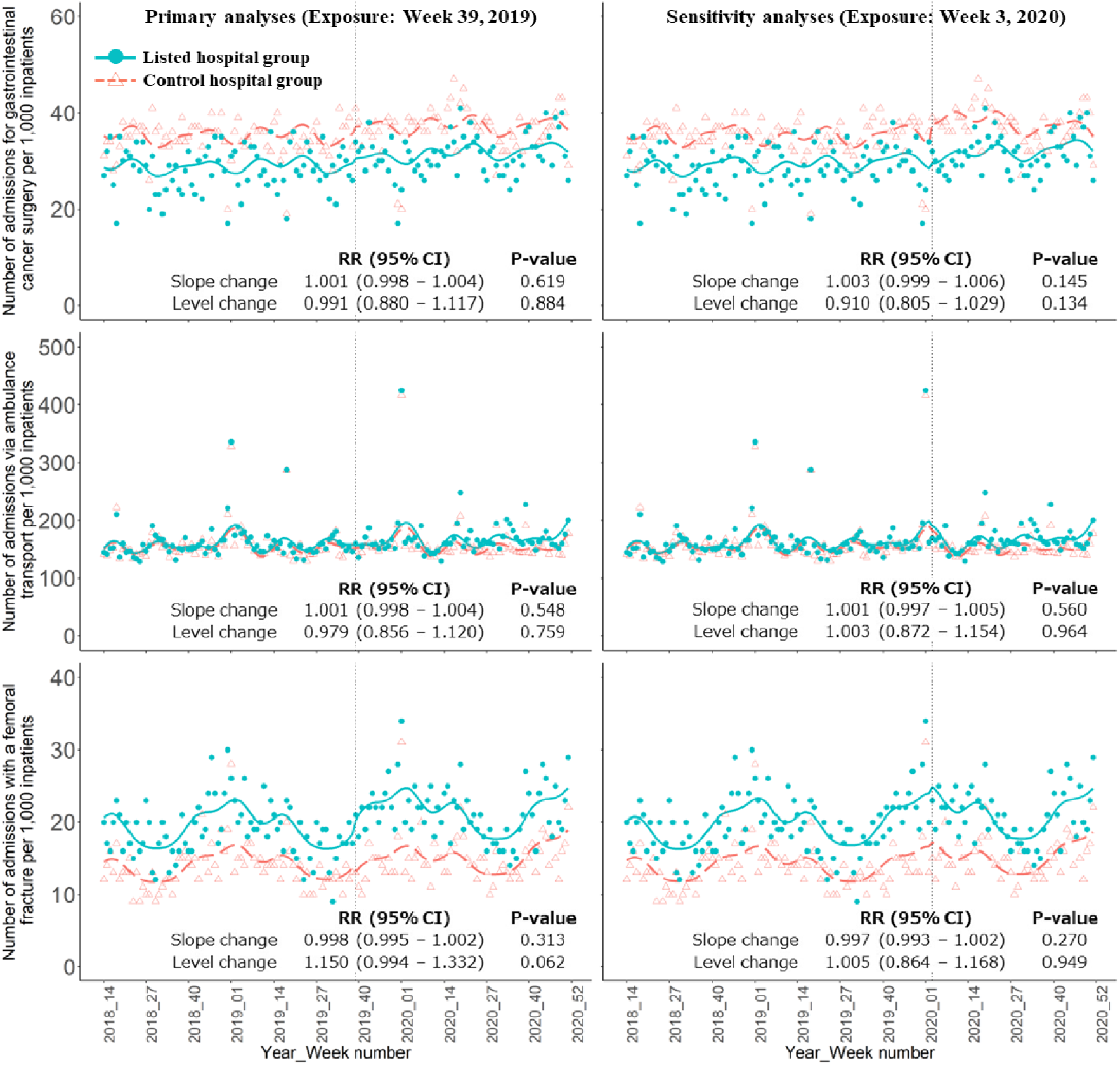
Weekly number of each admission cases per 1,000 newly admission cases with the results of CITS analysis. The three graphs on the left and right sides show the primary and sensitivity analysis results, respectively. The y-axis indicates the outcomes, and the x-axis indicates the year and week numbers. Each blue dot and orange triangle indicates the observed number for the listed and control hospital groups, respectively. Blue and dashed orange lines indicate the predicted trend in the listed and control hospital groups, respectively, based on the model. The vertical dashed line indicates the exposure. RR: Rate Ratio. CI: Confidence Interval.

## 4. Discussion

In this study, we showed the status of functional differentiation of hospital beds of the listed hospitals from FY 2019–2021 compared to non-listed hospitals using the bed functions report. Moreover, the impact of the release of the list on the number of three types of admission cases per 1,000 admissions at listed hospitals was evaluated by CITS analysis using DPC data.

The results of the first analysis revealed that the functional differentiation of hospital beds of listed hospitals differs from those of non-listed hospitals. The percentage of listed hospitals that decreased the total number of licensed beds or changed the function of licensed beds was higher than that of non-listed hospitals. In addition, the number of acute care beds in listed hospitals decreased more than in non-listed hospitals, while the number of convalescent beds increased more than in non-listed hospitals. Despite the changes in the listed hospitals being the same as in the MHLW report,^18^ this study revealed that the functional differentiation of hospital beds of the listed hospitals is more remarkable than the non-listed hospitals. In addition, although the report of MHLW reported that 44 hospitals did not intend to change their function,^18^ our analysis showed that restructuring the healthcare delivery system is progressing. Publication of the list in 2019 could be one reason for the differences between listed and non-listed hospitals. In addition, the Hospital Bed Functional Reorganization Support Program,^19, 27^ established in 2020, may also have encouraged hospitals to conduct functional differentiation of hospital beds, with a decrease in the total number of beds between 2019 and 2021.

The findings of the CITS analysis were that the release of the list had no impact on the number of the three types of admission cases at the listed hospital group compared to the control hospital group. Despite reputational concerns caused by the publication of the list and its being treated as a “hospital restructured list”,^21, 22, 36, 37^ including a hospital on the list did not affect the decisions of emergency services or patients in choosing where to transport or hospitalize. As Miyazawa indicated,^21^ many listed hospitals provided messages to patients and stakeholders through their websites and publications after the list was published, potentially counteracting the negative impact. Neither the primary nor sensitivity analyses showed a significant impact on the number of the three types of admission cases at the listed hospital group. It was suggested that the role-sharing related to the three types of diseases evaluated in this study in the listed hospital group was not implemented compared to the control hospital group or might be implemented regardless of the timing of the release of the list or the official notification from the MHLW. Although no impacts were apparent during this study period, impacts may be apparent in longer-term studies, and further research is needed to assess impacts over a longer period.

The restructuring of the healthcare delivery system in Japan is urgent. It is necessary to encourage discussions among public and private hospitals more actively. Policymakers must also be aware of the adverse effects of reorganization and integration. In the UK, the reduction in the number of beds has made it more difficult to adapt to fluctuations in demand.^38^ In Canada, overcrowding in emergency services rose during reorganization,^39^ and mortality rates increased in the group receiving intensive care after reducing the number of beds.^40^ Public and municipal hospitals in Japan play a key role in treating COVID-19 cases, especially in areas with fewer than 100,000 people.^41^ Implementing policies to promote reorganization and integration with the premise of continuing to protect community healthcare is desired based on an analysis of the impact of the list’s release on various perspectives. In addition, it is important to implement reorganization and integration policies based on the supply and demand of the number of beds and various medical resources. In Denmark, the Ministry of Health and its advisory body, the National Board of Health, assigned specialized physicians to different regions and succeeded in centralizing hospitals.^42^

This study has several limitations. First, we have not evaluated all aspects of hospital reorganization and integration; these generally include the change in the number of beds by medical function and the change in the hospitals; strategy without changing the number of beds.^43^ In this study, changes in the number of three types of cases were evaluated for reorganizations and consolidations that did not involve the number of beds. However, since this study selected and analyzed the three most important admission cases, for which many hospitals had already worked among Categories A and B, it is considered to have provided fundamental knowledge regarding the impact of the release of the list. Second, the list’s impact on smaller hospitals might have been more significant than on larger hospitals. In general, smaller hospitals tend to be more financially unstable^44^ and experience more pressure for reorganization and integration. Further research is needed to clarify the detailed impact of the release of the list on smaller hospitals, for example, through analyses that do not rely on DPC data.

Despite these limitations, this study is unique due to two perspectives. The first is that few studies have clarified the status of functional differentiation of hospital beds of listed hospitals compared to non-listed hospitals or have evaluated the impact of the release of the list, providing insight into the formulation of healthcare policy. Second, the CITS analysis, in which hospitals near the cutoff were used as the control hospital group, allowed us to compare whether there was an impact depending on whether the hospitals were classified as listed hospitals, which yielded reliable results.

## 5. Conclusions

Our analysis suggested that the listed hospitals underwent more significant functional differentiation of hospital beds than the non-listed hospitals, with a decrease in the number of acute care beds and an increase in convalescent beds. It also demonstrated that the release of the list did not influence the changes in the number of admissions for gastrointestinal cancer surgery, admission via ambulance, or for those with a femoral fracture per 1,000 admission cases respectively, in the listed hospital groups. in listed hospitals.

## Acknowledgments

We gratefully acknowledge the contributions of the staff members and all the participating hospitals.

## Appendix. The model and parameters in a controlled interrupted time series analysis, regarding the release of the list

The following model was used:

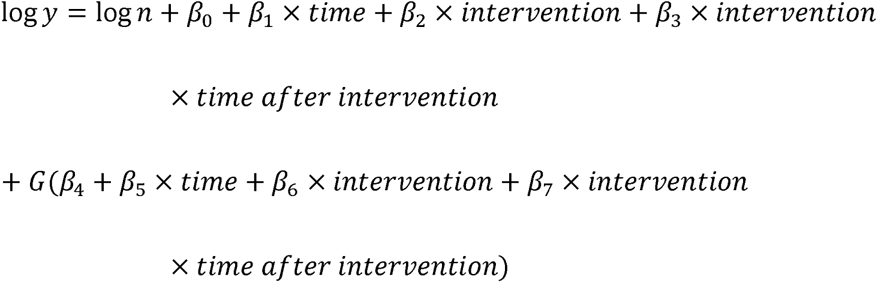

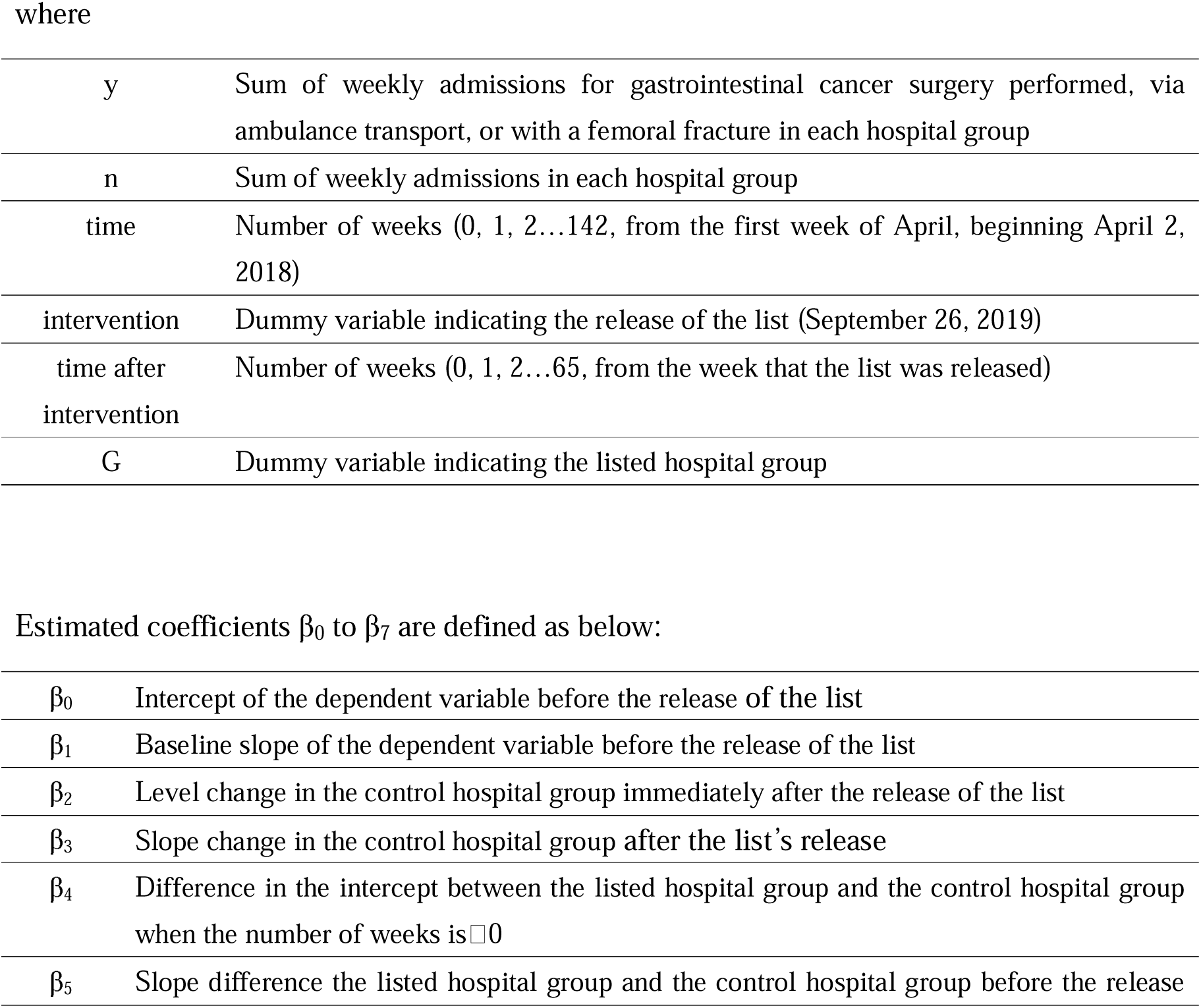
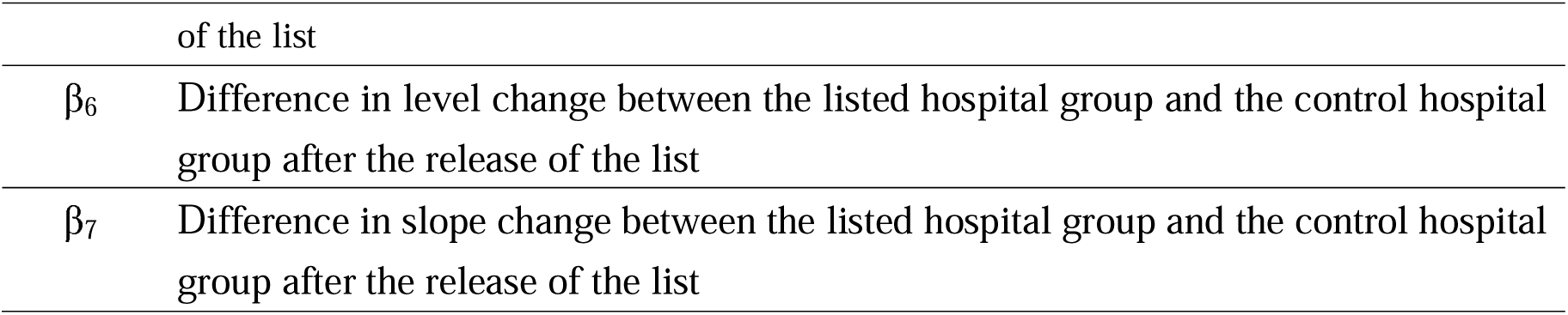

|  |  |
| --- | --- |
| y | Sum of weekly admissions for gastrointestinal cancer surgery performed, via ambulance transport, or with a femoral fracture in each hospital group |
| n | Sum of weekly admissions in each hospital group |
| time | Number of weeks (0, 1, 2...142, from the first week of April, beginning April 2, 2018) |
| intervention | Dummy variable indicating the release of the list (September 26, 2019) |
| time after intervention | Number of weeks (0, 1, 2...65, from the week that the list was released) |
| G | Dummy variable indicating the listed hospital group |
| Estimated coefficients $\beta_0$ to $\beta_7$ are defined as below: |  |
| $\beta_0$ | Intercept of the dependent variable before the release of the list |
| $\beta_1$ | Baseline slope of the dependent variable before the release of the list |
| $\beta_2$ | Level change in the control hospital group immediately after the release of the list |
| $\beta_3$ | Slope change in the control hospital group after the list's release |
| $\beta_4$ | Difference in the intercept between the listed hospital group and the control hospital group when the number of weeks is $\square 0$ |
| $\beta_5$ | Slope difference the listed hospital group and the control hospital group before the release |
|  | of the list |
| $\beta_6$ | Difference in level change between the listed hospital group and the control hospital group after the release of the list |
| $\beta_7$ | Difference in slope change between the listed hospital group and the control hospital group after the release of the list |

## Author contributions

Conceptualization: H.T., J.S, S.K., and Y.I.

Methodology: H.T., J.S., and Y.I.

Software: H.T., J.S, and S.K.

Validation: J.S, S.K., and Y.I.

Formal Analysis: H.T., J.S, and S.K.

Investigation: H.T., J.S, S.K., and Y.I.

Resources: S.K, and Y.I.

Data Curation: J.S., S.K., and K.F

Writing – Original Draft Preparation; H.T.

Writing – Review & Editing: All authors

Visualization: H.T

Supervision: Y.I.

Funding Acquisition: Y.I.

Project Administration: Y.I.

## Data Availability

According to the Ethical Guidelines for Medical and Health Research Involving Human Subjects of the MHLW, Japan (available from https://www.mhlw.go.jp/file/06-Seisakujouhou-10600000-Daijinkanboukouseikagakuka/0000 080278.pdf), providing information to study subjects is one of the necessary conditions for waiving informed consent. The information includes the range of data users. Therefore, the datasets generated and/or analyzed during this study are available from the corresponding author and the other contact points on reasonable request. The other contact points are the Office of Research Promotion, General Affairs and Planning Division, Kyoto University (; Tel: +81-75-753-9301) and the Ethics Committee, Graduate School of Medicine, Kyoto University.

