## Supplementary material for "How were Hospitals Affected by the Ministry’s Release of Hospital Names to be Potentially Reorganized?"

**Supplemental Table 1. Types of gastrointestinal cancer**

| Gastrointestinal cancer site | ICD-10 codes | Corresponding claims code |
|---|---|---|
| Esophageal | C15.0, C15.1, C15.2, C15.3, C15.4, C15.5, C15.8, C15.9 | K395, K508-2x, K522x, K522-x, K5261, K526-x, K527x, K529x, K529-2x, K529-3, K531x, K654 |
| Gastric | C16.x | K653x, K654-2, K654-3x, K6552, K655-22, K655-23, K655-42, K655-52, K655-53, K6572, K657-22, K657-23, K657-24, K6573 |
| Small bowel, peritoneal | C17.x, C26.8, C45.1, C48.0, C48.1, C48.2, C48.8, C77.2, C78.4 | K627x, K627-2x, K636, K636-3, K636-4, K643, K643-2, K653x, K654, K662, K662-2, K716x, K716-2x, K724, K726, K726-2 |
| Colon | C18.x, C26.0, C26.9, C78.5 | K6113, K627x, K627-2x, K636, K636-3, K636-4, K643, K643-2, K662, K662-2, K719-2x, K7193, K719-3, K721x, K721-4, K724, K732x, K736x |
| Rectal, anal | C19, C20, C21.x, C77.5 | K6113, K636, K636-3, K636-4, K645, K7193, K719-3, K721x, K721-4, K732x, K732-2, K736x, K739x, K739-x, K740x, K740-2x, K748x |
| Liver, intrahepatic cholangiocarcinoma | C22.x, C78.7 | K636, K636-3, K636-4, K677-2x, K695x, K695-2x, K697-2x, K697-3x |
| Gallbladder, extrahepatic bile duct | C23, C24.x | K672, K672-2, K675x, K677x, K677-2x, K703x, K695x, K695-2x |
| Pancreatic | C25.x, C26.1 | K700, K700-x, K702x, K702-2x, K703x, K704, K710, K711, K711-2 |

**Supplemental Figure 1. Flowchart of hospital classifications in this study**

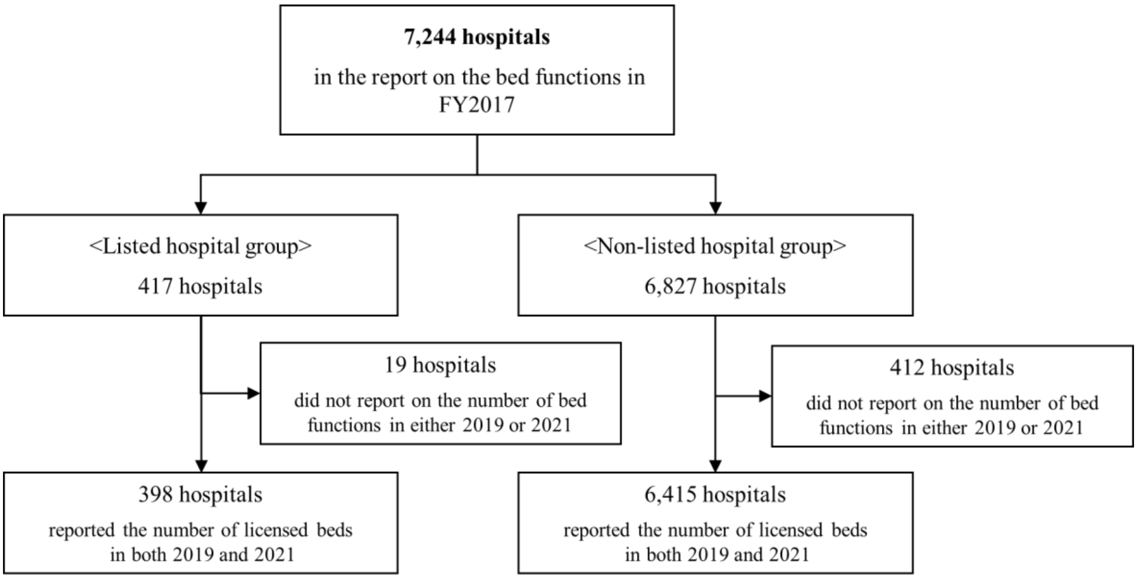
